# Effects of self-rated mental and physical work demands on cognition are dependent in a cross-sectional sample of the Health and Retirement Study

**DOI:** 10.1101/2024.11.11.24317122

**Authors:** Ruby C. Hickman, Herong Wang, Daniel J. Brandt, Erin B. Ware, Kelly M. Bakulski

## Abstract

**Objective:** This study assessed whether self-rated physical and mental work demands were associated with cognition among older working adults and whether their effects were dependent.

**Methods:** Our cross-sectional sample consisted of 6,377 working older adults uisng the Health and Retirement Study in 2004. Self-rated work demands were summarized from four questions about frequency of mental or physical demands in the respondent’s current job. Cognition was assessed using a subset of the Telephone Interview for Cognitive Status. We used multivariable linear regression to test for associations and additive interaction between physical and mental work demands and cognition, adjusted for age, sex, race, education, and practice effect.

**Results:** Independently, higher physical work demands were associated *(P*<0.001) with poorer cognition and higher mental work demands were associated (*P*<0.001) with better cognition. The effect of one work demand measure became more negative as level of the other increased (B for interaction = -0.23, 95% CI: -0.43, -0.03). A one-point increase in mental work demands was associated with 0.79 (95% CI: 0.51, 1.08) points higher cognition score when physical work demands were lowest, but was not associated with cognition when physical work demands were highest (0.11, 95% CI: -0.26, 0.48). The highest predicted cognition score was for the highest mental and lowest physical work demands. Results were robust to additional adjustment for health and behavior covariates.

**Conclusions:** The associations of self-rated mental and physical work demands on cognition are dependent. Future studies should strongly consider examining interactions to capture the range of work demand effects.

**Key points:** *What is already known on this topic:* Mental work demands may help to preserve cognition in older adults, both while working and after retirement. However, the relationship of physical work demands and its interaction with mental work demands on cognition is less clear.

*What this study adds:* We found that the associations of mental and physical work demands on cognition are dependent. There was a statistically significant interaction between mental and physical work demands, with the beneficial effect of mental work demands attenuating with increasing levels of physical work demands.

*How this study might affect research, practice or policy:* This study suggested the interactive impact of mental and physical work demands on cognition. Future studies examining work demands need to consider interaction between mental and physical work demands. More generally, this means that the traditional view of mentally stimulating work as protective of cognition may not apply for workers who also have physically demanding jobs. The effects that our jobs have on our cognitive health is complex, and ever more important as we live and work longer.

## Introduction

As the US population ages, the number of older adults in the workforce has increased. From 1998 to 2018, the proportion of the labor force aged 55 or older nearly doubled, from 12.4% to 23.1% (Clark & Ritter, 2020). The aging population also means that more people are affected by cognitive impairment and dementia. As of 2022, an estimated 6.5 million Americans age 65 or older live with Alzheimer’s dementia (“2022 Alzheimer’s Disease Facts and Figures,” 2022). Cognitive impairment and cognitive decline affect even more people, with the worldwide prevalence of mild cognitive impairment estimated to be between 15-20% among persons 60 years and above (Petersen, 2016). As more older adults remain in the workforce longer, it becomes even more important to understand the relationship between work characteristics and cognition.

Mental work demands may help to preserve cognition in older adults, both while working and after retirement. Higher mental work demands based on job title are associated with reduced odds of dementia later in life (Andel et al., 2005; Kröger et al., 2008; Smyth et al., 2004; Then et al., 2017) and improved cognition (Fisher et al., 2014; Potter et al., 2006), including in younger adults (Nexø et al., 2016). Higher self-rated mental work demands are also associated with reduced risk of cognitive impairment (Bosma et al., 2003). While the exact mechanism is not understood, evidence suggests that increased mental work demands are associated with improved cognitive outcomes.

The relationship between physical work demands and cognition is less clear. Higher levels of leisure-time physical activity are consistently associated with improved cognition (Ludyga et al., 2020). Conversely, higher occupational physical demands are associated with poorer cognition-related outcomes. Increased occupational physical demands based on job title are associated with worse cognition-related outcomes, including increased odds of Alzheimer’s disease (Smyth et al., 2004) and poorer longitudinal cognition trends (Potter et al., 2006). In the U.S. Health and Retirement Study (HRS), physical demands assigned by job title were associated with lower baseline cognition, but also marginally associated with slower cognitive decline (Lee et al., 2022). Self-rated occupational physical demands were not associated with risk of dementia or Alzheimer’s disease (Rovio et al., 2007) or odds of cognitive impairment (Ku et al., 2012), but were associated with reduced incident cognitive impairment non-dementia (Marengoni et al., 2011) and with poorer cognition in the 2010 wave of the Health and Retirement study (Choi et al., 2022).

Several studies have examined the effects of both mental and physical work demands on cognition- or dementia-related outcomes (Potter et al., 2006; Prieske et al., 2021; Smyth et al., 2004), but only one study has examined their effects in the same model, allowing researchers to examine the effect of one work demand measure while holding the level of the other constant. In a study of Canadian older adults using work demands assigned by job title, complexity of work with people and things, but not data, was associated with reduced hazard of dementia when adjusted for work-related physical activity (Kröger et al., 2008). To our knowledge, this is one of the first studies that has examined the presence of interaction between physical and mental work demands and included self-rated physical and mental work demands in the same model. Self-rated work demands, as opposed to demands assigned by job title, may better capture the range of work demands actually experienced by participants.

The aims of this study are to test whether self-rated physical and mental work demands are associated with cognition among older working adults and how their effects may be dependent. Using data from the Health and Retirement Study, a longitudinal panel study of U.S. adults over the age of 50, we examine the relationship between self-rated physical work demands, self-rated mental work demands, and cognition, as well as the presence of an additive interaction between self-rated mental and physical demands. This study will advance our understanding of how physical and mental work demands, two co-occurring exposures, may jointly affect cognition among older adults.

## Methods

### Study sample

The Health and Retirement Study (HRS) is a longitudinal panel study of U.S. adults 50 years and older and their spouses that began in 1992 and is conducted by the University of Michigan and funded by National Institute on Aging (NIA U01AG009740). This secondary analysis was approved by the University of Michigan Institutional Review Board (HUM00128220). Participants provided written informed consent. HRS participants are surveyed biennially on a range of topics including income and wealth, health, work and retirement, and family connections (Sonnega et al., 2014). Participants in the 2004 wave of the HRS were included in these analyses. We chose to analyze the 2004 wave for two primary reasons. First, a new cohort of participants, the Early Baby Boomers (born 1948-53), was added in 2004, increasing the number of younger and working participants. Second, work characteristics were asked of all participants in the 2002 panel and of new participants in the 2004 panel. This is not the case in future waves, where values for most work demand measures were carried forward unless a participant changed jobs, resulting in exposure measures which may be more inaccurate for participants who had stayed in the same job or been in the study longer.

### Data sources

All survey measures were collected as part of the HRS core survey and are publicly available. We used the HRS imputation of cognitive functioning measures, details of which are available here (McCammon et al., 2023). If available, data was pulled from the RAND corporation longitudinal dataset (Health and Retirement Study, 2024; RAND HRS Longitudinal File 2020 (V2), 2024), which is a cleaned data product containing information on HRS variables. If not available in the longitudinal file, variables were pulled from the RAND Fat files, which presents the survey data in a single respondent-level dataset for each wave (Health and Retirement Study, 2017; Health and Retirement Study, 2019; RAND HRS 2002 Fat File (V2C), 2017; RAND HRS 2004 Fat File (V1C), 2019).

### Cognitive outcome

Cognition was assessed in 2004 using an adapted version of the Telephone Interview for Cognitive Status, which assesses two separate aspects of cognition: episodic memory and mental status. Episodic memory was assessed by asking participants to recall a list of 10 words immediately and after a 5 minute delay resulting in a score of 0-20 (one point per word recalled at each time point). Mental status was assessed through a serial seven subtraction and backward count from 20 tasks resulting in a score of 0-7, with 5 points possible from the serial 7 subtraction (one point per correct subtraction), and 2 points for backward count from 20 (two if correct on first try, one if correct on second try, zero for two failed attempts).

Our main outcome was the combined score of the two subscales, which has been previously validated in HRS to classify respondents into normal cognition, cognitive impairment non-dementia (CIND), and dementia (Crimmins et al., 2011) using score ranges of 12-27, 7-11, and 0-6, respectively. To increase statistical precision and to expand our inference to the spectrum of cognition beyond impairment, we used the continuous score, ranging from 0 to 27, as our primary outcome measure. In sensitivity analyses, we considered CIND/dementia vs. normal cognition, the episodic memory subscale (20 points total), and the mental status subscale (7 points total) as alternate outcomes.

### Exposure assessment

Work demands were assessed via a series of Likert scale assessments, which we transformed into summary scores. The physical work demands summary score was derived from four items asking about the frequency of their job requiring “lots of physical effort,” “stooping, kneeling, or crouching,” “good eyesight,” and “lifting heavy loads,” from the following options: none or almost none of the time, some of the time, most of the time, and all or almost all the time. Each question was scored from 0-3 respectively, and responses were averaged across the four physical work demands questions to create a summary score ranging from 0-3, with 0 indicating the lowest and 3 representing the greatest amount of physical work demands.

Mental work demands were assessed via four questions. The first three, similar to that of physical work demands, asked about the frequency of their job requiring “intense concentration,” “skill in dealing with people,” and “work with computers,” from the following options: none or almost none of the time, some of the time, most of the time, and all or almost all of the time. The last question asked about agreement with the statement “my job requires me to do more difficult things than it used to” with the following options: strongly disagree, disagree, agree, or strongly agree. Each of the four items was assigned a score from 0-3 and averaged to create a summary score ranging from 0-3, with 0 indicating the lowest amount, and 3 representing the greatest amount of mental work demands.

Work demand measures were carried forward from the 2002 wave and not asked again in 2004 except in the following situations. Participants who joined the study or changed jobs in 2004 were asked all work demand measures in 2004. Additionally, all participants answered questions on “computer use” and “job requiring more difficult things than it used to” in 2004 regardless of job tenure.

### Covariate measures

Respondents’ age in years was assessed at administration of the 2004 questionnaire. Sex (male or female), years of education, and race/ethnicity was assessed upon entry into HRS. For race/ethnicity, respondents were classified into non-Hispanic White, non-Hispanic Black, and other. Cognitive test practice effect was defined as present when the participant had experience taking the same test in any previous wave of HRS. Job tenure was length of current job in years. Household, for the purpose of this analysis, is the combination of a participant’s original household assignment upon entry to HRS, and their sub-household assignment, which was assigned in case of household split or merge (e.g., a participant divorces, remarries, etc.). The combined household ID allowed us to assess and account for similarities between current household members.

Depressive symptoms was a dichotomous variable using an 8-item short-form of the Center for Epidemiologic Studies Depression Scale (CESD) (Radloff, 1977). A score of 0 indicated no symptoms and a score of 1-8 indicated any symptoms. Smoking status was classified as never, former, or current smoker using ever and current smoker variables from RAND. We obtained diabetes, hypertension, and stroke history from the participant’s self-report of a diagnosis in any wave of the study up to 2004. Drinks per week was calculated from participant report of average drinks per day and average days drinking per week. Participants rated their health as poor, fair, good, very good, or excellent. We merged poor and fair due to low frequency of poor health. We used three leisure-time physical activity variables that assessed frequency (every day, > once per week, once per week, 1-3 times per month, rarely or never) of mild, moderate, and vigorous activity. We combined “every day” and “> once per week” due to low frequencies and because “every day” was not an option given by interviewers and was only recorded if participants volunteered it.

### Statistical methods

All analyses were completed using R (version 4.0.2) (R Core Team, 2020). We tested differences in covariate distributions between the following groups using t-tests or ANOVA for continuous variables and chi-square tests for overall association of categorical variables. First, we compared the included and excluded sample. Second, we compared those in each of the four combinations of mental and physical work demands below and above or equal to the median (< 2 vs. ≥ 2 for mental work demands, < 1.25 vs ≥ 1.25 for physical work demands). Third, we compared those with impaired (Langa-Weir classification of cognitive impairment no dementia or dementia, cognition score 0-11) and non-impaired (cognition score 12-27) cognition. To test intra-group correlations within household that could affect independence of our linear regression models, we used Pearson’s correlation coefficient. There were few (0.6%) participants living in households with >2 members, so for the purpose of simplicity, we limited this analysis to households with 2 members.

We used multiple linear regression to estimate the associations between physical work demands, mental work demands, and cognition. We examined each exposure independently in unadjusted models, then included both exposures in the same model for all subsequent analyses. We examined the crude and adjusted associations, adjusting for age, sex, race/ethnicity, years of education, and practice effect. We tested for additive interaction between physical and mental work demands.

We tested robustness to potential confounders adding two distinct sets of covariates. First, we added health-related measures (depressive symptoms, hypertension, diabetes, self-rated health, and stroke), and second, we added health behavior measures (smoking, drinking, frequency of mild, moderate, and vigorous physical activity). We assessed robustness to violation of the independence assumption from household clustering using a mixed effects model with a random intercept for household using the lme4 package (Bates et al., 2015) and calculating the intra-class correlation coefficient (ICC). We used Cook’s distance and a visual assessment to identify potential influential points and removed them in a sensitivity analysis.

We tested the consistency of our results when using alternate cognitive outcome measures. We used logistic regression to estimate the association of mental and physical work demands with the odds of impaired (cognition score 0-11) vs. normal cognition (score 12-27). For the subscale analyses, we used linear regression to estimate the association between mental and physical work demands and score on the episodic memory subscale (delayed and immediate recall). The mental status subscale (range 0-7) was highly left-skewed, so we dichotomized it at the median (7) and used logistic regression to estimate the association of mental and physical work demands with the odds of a mental status subscale (range 0-7) score at/above or below the median (7 vs. <7).

## Results

### Study sample characteristics

Of the 7,841 participants worked for pay in the 2004 wave, we excluded 635 (8.1%) participants under age 50. A further 829 (10.6%) of participants were excluded for missing data (**Supplemental Figure 1**) leaving our analytic sample at 6,377.

Participants included in our analysis had an average age of 60, were 46% male, 75% non-Hispanic White, and had completed an average of 13.4 years of education **(Supplemental Table 1)**. The average cognition score was 16.8 points out of 27 total possible points, and 68% had practice effect, meaning they had taken the same test in a previous wave of the study. Included participants were similar to excluded participants in cognition classification and alcohol consumption; however, included participants were more likely to have lower physical and mental work demands, be non-Hispanic White, have more years of education, and be non-current smokers compared to excluded participants.

### Descriptive statistics and bivariate measures by joint exposure level

For our descriptive statistic comparison, we classified participants into mental and physical work demand groups based on the median values in our sample (i.e. low mental/low physical, low mental/high physical, high mental/low physical, high mental/high physical demands). Mean cognition score was highest (mean = 18.0, standard deviation (SD) = 3.2), in participants who had high mental work demands (above or equal to the median ≥ 2) and low physical work demands (below the median <1.25). Mean cognition score was lowest (mean = 15.7, SD = 3.8) among those with low mental work demands and high physical work demands **(Table 1)**. Education was also highest in those with high mental and low physical work demands (mean = 14.6 years, SD = 2.2), and lowest in those with low mental and high physical demands (mean = 12.1 years, SD = 2.9). The distribution of race/ethnicity, age, sex, and other covariates differed between the different work demands groups.

**Table 1.**
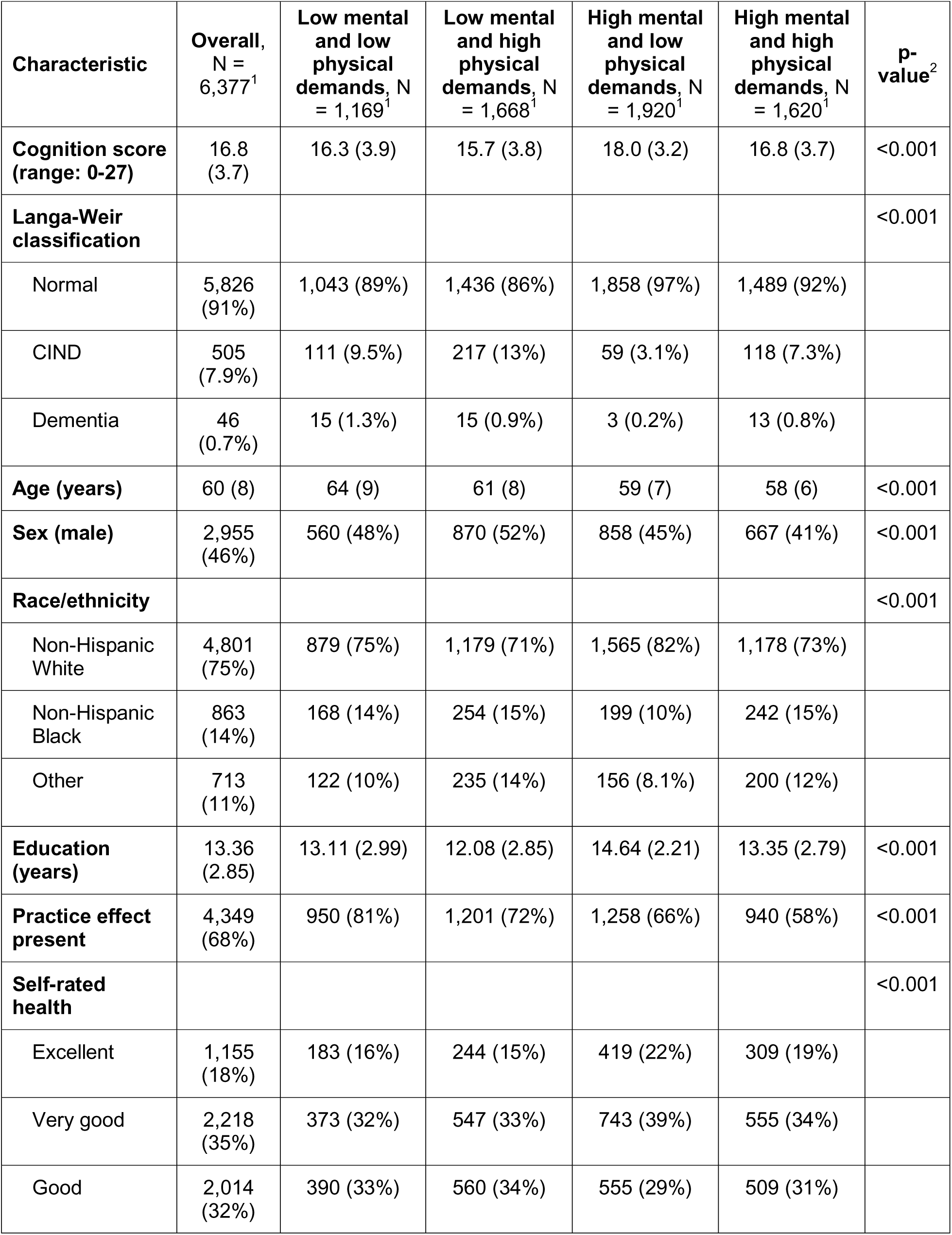
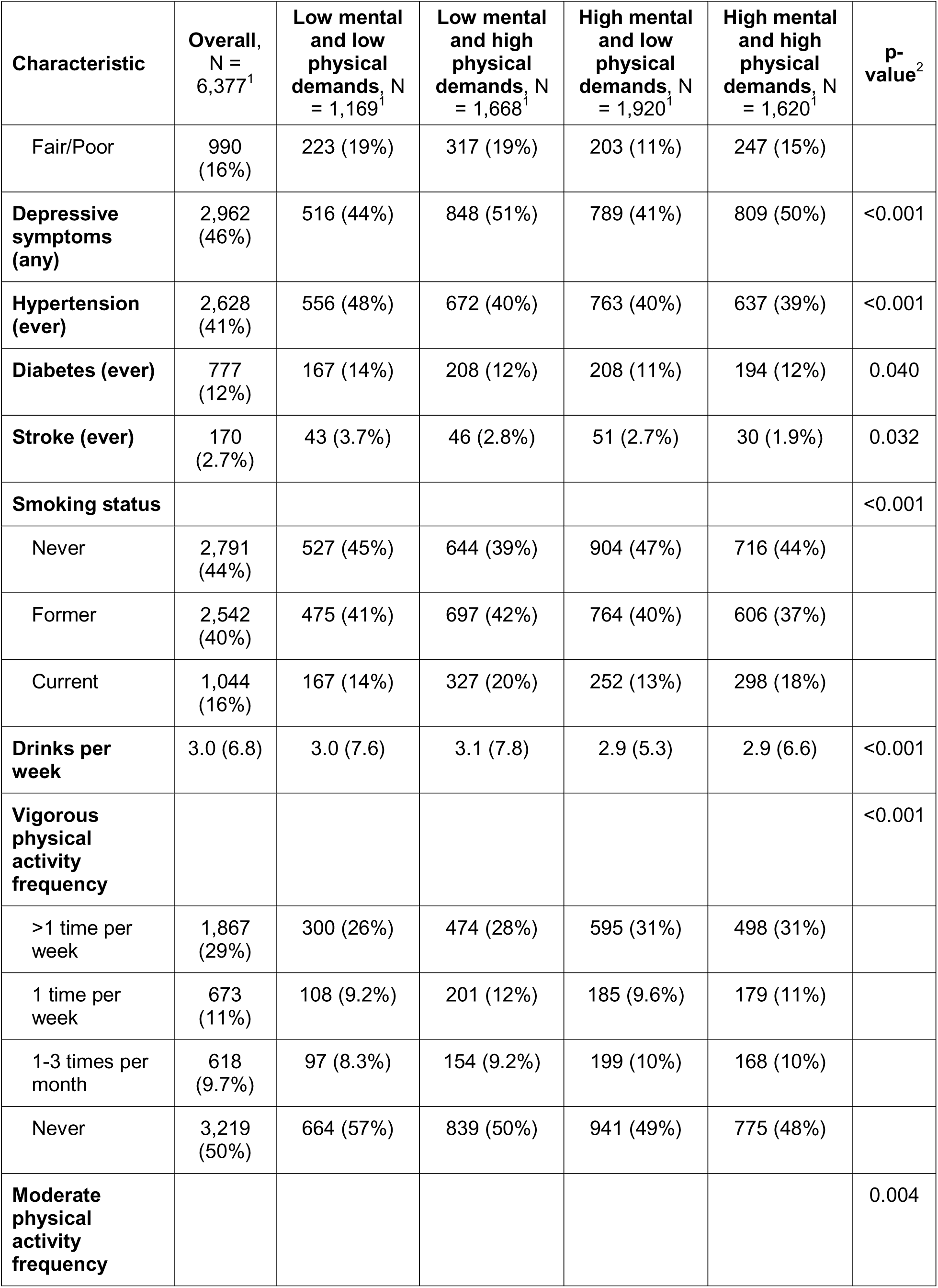
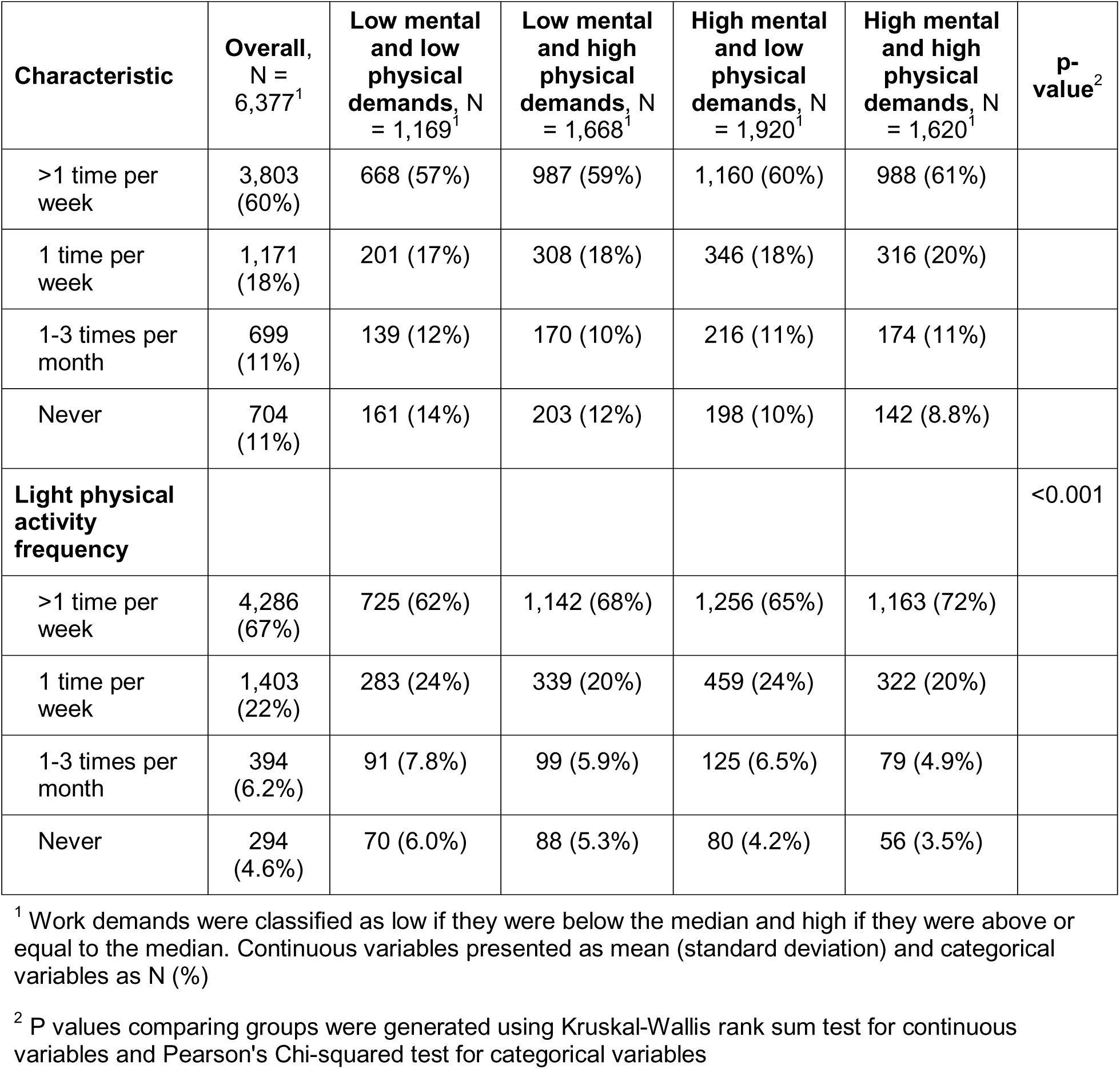
Characteristics of included study participants by joint exposure level, Health and Retirement Study, 2004 wave.

Participants who were classified as having impaired cognition (cognition score 0-11, n=551) were older, more likely to be non-White, and had completed fewer years of education than participants with normal cognition (n=5,826) (**Supplemental Table 2**). Mental work demand score was lower (1.68 vs. 1.99*, P* <0.001), and the physical work demand score was higher (1.54 vs 1.29, *P* <0.001) in those with impaired cognition than those with normal cognition. More participants with impaired cognition were depressed, were current smokers, drunk more, had ever been diagnosed with hypertension, diabetes, or stroke, and rated their health as good or fair/poor. There was no difference in sex, or practice effect between those with impaired or non-impaired cognition.

### Primary regression analyses

Self-rated mental work demand score was slightly negatively correlated (r = -0.09, *P*<0.001) with self-rated physical work demand score. In an unadjusted model, a one-unit increase in self-rated mental work demands was associated with a 1.39 increase in cognition score (95% CI 1.25, 1.54) (**Table 2**). In a separate unadjusted model, a one-unit increase in level of self-rated physical work demands was associated with -0.88 point decrease in cognition score (95% CI -1.00, -0.75). When included in the same model with no interaction term, the independent effects of mental and physical work demands were similar in magnitude and direction and remained significant. Adding age, sex, race/ethnicity, years of education, and practice effect to this co-adjusted model attenuated the effects of mental work demands (change per one unit increase: 0.50, 95% CI 0.36, 0.65) and the effect of physical work demands (change per one unit increase: -0.31, 95% CI -0.44, -0.19) though they remained significant. The effect of a 1-unit increase in mental work demands controlling for level of physical work demands was similar in magnitude to the negative effect of a 5-year increase in age (-0.45), indicating that there is a meaningful association between mental work demands and cognition.

**Table 2.** Linear regression associations between self-rated physical and mental work demands and cognition score, Health and Retirement Study, 2004 wave.

| Models | Parameter (B, 95% CI) |  |  | Adjusted R<sup>2</sup> |
| --- | --- | --- | --- | --- |
|  | Mental work demands | Physical work demands | Mental*Physical work demands |  |
| **Individual unadjusted<sup>1</sup>** | 1.39 (1.25, 1.54) | -0.88 (-1.00, -0.75) | --- |  |
| **Joint unadjusted<sup>2</sup>** | 1.31 (1.17, 1.45) | -0.77 (-0.90, -0.65) | --- | 0.07 |
| **Demographic<sup>3</sup>** | 0.50 (0.36, 0.65) | -0.31 (-0.44, -0.19) | --- | 0.21 |
| **Demographic + interaction<sup>4</sup>** | 0.79 (0.51, 1.08) | 0.12 (-0.27, 0.52) | -0.23 (-0.43, -0.03) | 0.21 |
| **Health sensitivity<sup>5</sup>** | 0.72 (0.44, 1.01) | 0.06 (-0.33, 0.45) | -0.19 (-0.38, 0.01) | 0.23 |
| **Behavior sensitivity<sup>6</sup>** | 0.79 (0.51, 1.08) | 0.08 (-0.31, 0.48) | -0.22 (-0.41, -0.02) | 0.22 |
| **Health + behavior sensitivity<sup>7</sup>** | 0.72 (0.44, 1.01) | 0.03 (-0.36, 0.42) | -0.18 (-0.38, 0.01) | 0.23 |

<sup>1</sup> only includes mental or physical work demand
<sup>2</sup> includes both mental and physical work demand
<sup>3</sup> adds age, sex, race/ethnicity, years of education, and practice effect to joint unadjusted model
<sup>4</sup> adds interaction term for mental\*physical work demand to demographic model
<sup>5</sup> adds depressive symptoms, self-rated health, hypertension, diabetes, stroke, and interaction term for mental\*physical work demand to demographic model
<sup>6</sup> adds smoking, drinking, physical activity, and interaction term for mental\*physical work demand to demographic model
<sup>7</sup> includes all parameters from health and behavior sensitivity models.

In the primary interaction model with age, sex, race/ethnicity, education, and practice effect as covariates, there was a significant additive interaction between mental and physical work demands (*P* = 0.02). Specifically, for each increasing unit of one of the work demand measures, the effect of the other on cognition became more negative (β for interaction: -0.23, 95% CI: -0.43, -0.03). For instance, when the physical work demands score was zero, the estimated effect of a one-unit increase in mental work demands was a gain of 0.79 (95% CI: 0.51, 1.08) points on the cognition test. However, at the highest level of physical work demands (three) the estimated effect of a one-unit increase in mental work demands was reduced to a gain of 0.11 (95% CI: -0.26, 0.48) points, holding all other variables constant. When mental work demands are zero, the estimated effect of physical work demands was slightly positive, though not significantly, (change per one unit increase: 0.12, 95% CI: -0.27, 0.52) on cognition score. At the highest level of mental work demands (three), a one-unit increase in physical work demands was associated with -0.57 (95% CI: -0.81, -0.32) points on the cognition score. Predicted cognition scores for all combinations of mental and physical work demand levels with covariates set at the mean or reference values are represented by the shading in **Figure 1**, showing that the highest predicted cognition score is for those with the highest level of mental work demands and lowest level of physical work demands.

**Figure 1.**
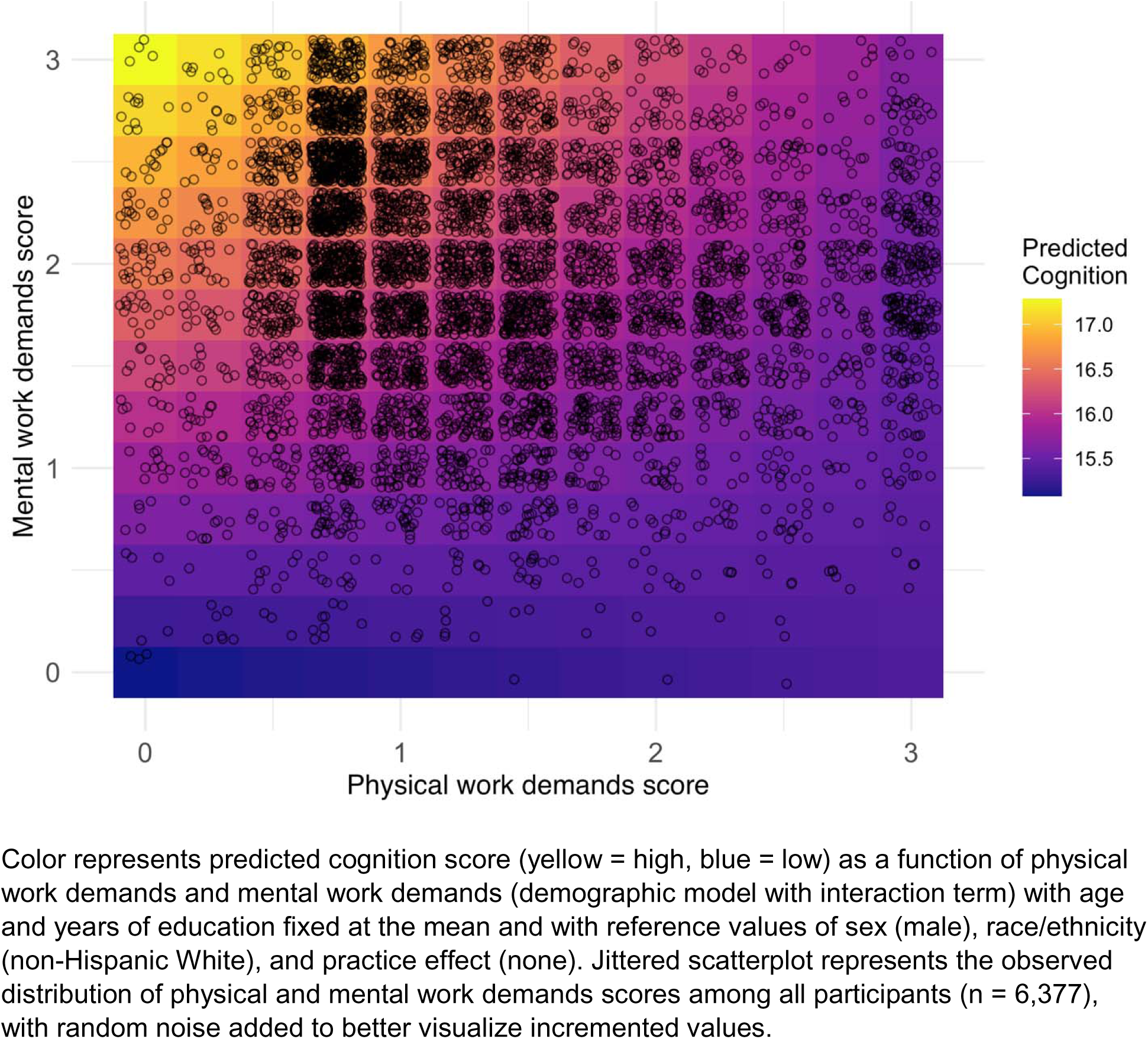
Surface of predicted cognition score as a function of self-rated physical work demands and self-rated mental work demands in the Health and Retirement Study from the 2004 wave.

### Sensitivity analyses

To assess the robustness of our findings, we performed a number of sensitivity analyses. We additionally adjusted our primary interaction model for a set of health-related potential confounders (self-rated health, depression, hypertension, stroke and diabetes), a set of behavior-related potential confounders (smoking, drinking, and physical activity), and both of them separately. We observed effect estimates that were similar in direction and significance to the primary interaction model (**Table 2**).

HRS samples spouses of participants, so to test if the independence assumption of linear regression is violated when we examined clustering by household. In our sample, 2286 (35.9%) of participants were part of a household with at least one other participant. Participants living in the same household were generally more similar on characteristics than would be expected at random. Participants in the same household were similar on cognition (r = 0.16, *P* <0.001), physical work demands (r = 0.17, *P* <0.001), and mental work demands (r = 0.19, *P* <0.001). However, results from a mixed effects model were nearly identical to our primary model (results not shown), and the ICC was low (0.02) indicating that the impact of household clustering was minimal.

In our primary interaction model, linear regression diagnostics suggested assumptions were met, with normally distributed residuals and homoscedasticity. However, predicted cognition scores were left skewed, with few individuals having a low predicted cognition score. While none of these observations exceeded Cook’s distance, a marker of influential outliers, we conducted a sensitivity analysis removing points in the bottom 20% of the range of predicted cognition values (n = 33) to test the robustness of our effect estimates. The impact of removing these observations on did not change the magnitude or interpretations of both main work demands effects and interaction effect (**Table 3).**

**Table 3.**
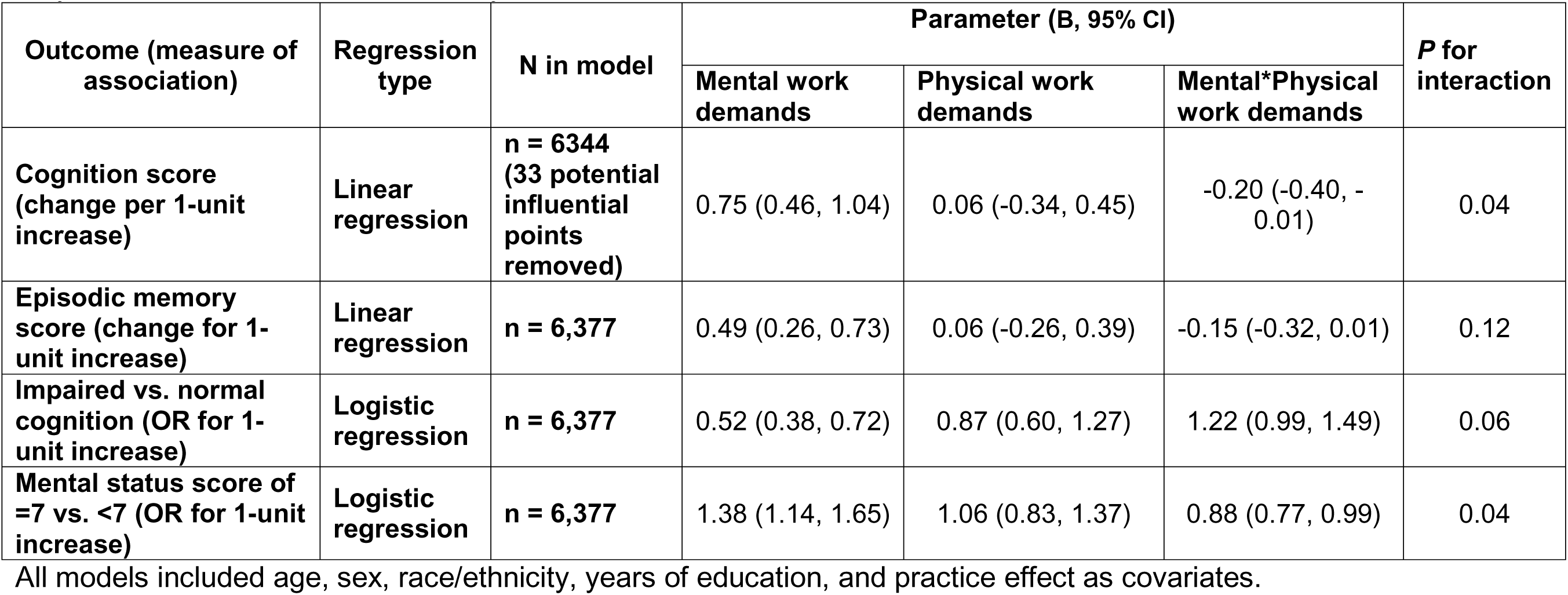
Associations between self-rated physical and mental work demands and several outcomes from sensitivity analyses, Health and Retirement Study, 2004 wave.

| Outcome (measure of association) | Regression type | N in model | Parameter (B, 95% CI) |  |  | P for interaction |
| --- | --- | --- | --- | --- | --- | --- |
|  |  |  | Mental work demands | Physical work demands | Mental*Physical work demands |  |
| Cognition score (change per 1-unit increase) | Linear regression | n = 6344 (33 potential influential points removed) | 0.75 (0.46, 1.04) | 0.06 (-0.34, 0.45) | -0.20 (-0.40, -0.01) | 0.04 |
| Episodic memory score (change for 1-unit increase) | Linear regression | n = 6,377 | 0.49 (0.26, 0.73) | 0.06 (-0.26, 0.39) | -0.15 (-0.32, 0.01) | 0.12 |
| Impaired vs. normal cognition (OR for 1-unit increase) | Logistic regression | n = 6,377 | 0.52 (0.38, 0.72) | 0.87 (0.60, 1.27) | 1.22 (0.99, 1.49) | 0.06 |
| Mental status score of =7 vs. <7 (OR for 1-unit increase) | Logistic regression | n = 6,377 | 1.38 (1.14, 1.65) | 1.06 (0.83, 1.37) | 0.88 (0.77, 0.99) | 0.04 |

All models included age, sex, race/ethnicity, years of education, and practice effect as covariates.

To assess clinical relevance, we used a dichotomous cognitive outcome (cognitively impaired versus not impaired), modeled with logistic regression and adjusted for the same covariates as our primary interaction model. The directions of effect for mental work demands, physical work demands, and the interaction term were the same as in our main analysis, though the interaction term became marginally significant (*P* = 0.06). The interaction term (odds ratio (OR) = 1.22, 95% CI: 0.99, 1.49) indicated that for increasing level of one work demand measure, the OR of cognitive impairment for the other will become more positive. For example, when physical work demands was zero, the effect of a one-unit increase in mental work demands was a 48% reduction in the odds of impaired cognition (OR = 0.52, 95% CI = 0.38, 0.72). However, with the highest level of physical work demands (3), the effect of a one-unit increase was a 5% reduction in the odds, an effect that was non-significant (OR = 0.95, 95% CI: 0.66, 1.38).

To investigate cognitive domains within our summary cognitive measure, cognitive subscales were used as outcomes. Using continuous episodic memory (immediate and delayed recall) as an outcome, we observed similar directions and levels of significance on the effect estimates of work demand to the main analysis using the entire 27-point scale. Using logistic regression to model the odds of a mental status score at/above vs. below the median (7 versus <7). We found a significant interaction (*P* = 0.04), and similar directions of effect to the main analysis. Together, in all sensitivity models, though the level of significance varied, we observed a beneficial main effect of increased mental work demands on cognition, a non-significant main effect for physical work demands, and an interaction term that indicated a more harmful (or less beneficial) effect of one work demand measure when the level of the other increased.

## Discussion

This study investigated the relationship between self-rated mental work demands, self-rated physical work demands, and their interaction between work demand measures on cognition in working older adults in the U.S. Independently, greater mental work demands were associated with higher levels cognition and greater physical work demands were associated with poorer cognition. In an interaction analysis, there was a statistically significant interaction between mental and physical work demands, with the beneficial effect of mental work demands attenuating with increasing levels of physical work demands. Results were robust after adjusting for potential health or behavior-related confounders and removal of potential influential points. When assessing the relationship between mental and physical work demands with related outcomes, directions of effects were similar.

While many studies have examined independent effects of mental and physical work demands on cognition, no study has reported on the presence of an interaction between these work characteristics. Our study supports previously reported findings that independently, mental work demands were meaningfully positively associated with cognition and physical work demands were negatively associated with cognition. The associations between mental and physical work demands and cognition were similar in a co-adjusted analysis, which reflects the findings of Kroger et al. that the reduced hazard of dementia associated with complexity of work with people and things was unchanged by controlling for work-related physical activity (Kröger et al., 2008). However, the interaction analysis suggests that their effects are dependent. This dependency means that while there is a strong body of literature supporting a positive effect of mental work demands, this effect may be significantly attenuated for workers who also have a physically demanding job.

The protective association between mental work demands or occupational complexity is often explained using the concept of cognitive reserve (Fisher et al., 2017; Pettigrew & Soldan, 2019). Cognitive reserve, as defined by a 2018 consensus paper, is the “adaptability that helps to explain differential susceptibility of cognitive abilities or day-to-day function to brain aging, pathology, or insult” (Stern et al., 2020). The results of our analysis suggest that increased cognitive reserve through higher mental work demands may not protect from potential insults from physical work demands, as evidenced by the significantly attenuated effect of mental work demands as level of physical work demands increase.

Physical work demands were negatively associated with cognition at all but the lowest level of mental work demands. There are several hypotheses that could potentially explain these effects (Fisher et al., 2017). First, physically demanding jobs are proxies for other variables including socio-economic status, education, and income that also are associated with cognition. Second, physically demanding jobs may be repetitive in nature and lack complex and varied intellectual demands. Our study attempted to address both of these through controlling for education and for mental work demands. A third hypothesis suggests a causal pathway for physical work demands through injury and inflammation from repetitive movement. Results from our study support a negative association between physical work demands and cognition for participants who reported non-zero mental work demands, which could potentially be explained by injury or inflammation.

In sensitivity analyses with alternate outcomes (impaired vs. non-impaired cognition, episodic memory subscale, and mental status subscale), directions of effects remained the same. The interaction effect was not significant for the more clinically oriented outcome of impaired vs. normal cognition, which could potentially be due to the loss of power from dichotomizing the outcome.

Our investigation has several strengths. It is the first study to examine interaction between physical and mental work demands on cognition, and our findings suggest a significant and meaningful interaction. The Health and Retirement Study is a large and diverse cohort that allows us to examine the effects of mental and physical work demands for older adults who have a wide range of jobs, education levels, and health statuses. Our use of self-rated work demands may better capture the actual work demands experienced by each individual rather than linking to an occupational database such as the Occupational Information Network (O*NET), and our consistency with previous results suggest that they are comparable. Analyzing a continuous cognitive outcome can increase power to capture differences that may not show on a clinical level. Our results were robust to a variety of potential confounders as well as removal of potential influential observations.

The greatest limitations of our study stem from the cross-sectional analysis. It is impossible to rule out reverse causation, so it could be that individuals who have worse cognitive function consequently self-select into jobs with lower mental work demands or higher physical work demands. Job tenure did not differ between participants with impaired vs. normal cognition, suggesting that individuals are not changing jobs differentially by cognition status. However, it is possible that individuals have adjusted work demands within a position depending on cognition level. Our study also could be prone to selection bias, as we selected only current workers, who are more likely to be healthy than the non-working population. Investigators in the future should pursue longitudinal analyses to address both direction of causation and selection bias, by collecting job demands at a past time and examining later cognition or cognitive trajectories.

## Conclusion

This study has broad implications for the impact of work demands on cognition. Future studies examining work demands need to consider interaction between mental and physical work demands. More generally, this means that the traditional view of mentally stimulating work as protective of cognition may not apply for workers who also have physically demanding jobs, such as nurses or first responders. The effects that our jobs have on our cognitive health is complex, and ever more important as we live and work longer.

## Data Availability

The Health and Retirement Study data used in this analysis are publicly available (https://hrsdata.isr.umich.edu/). The code used to process the data and produce the analyses are publicly available (https://github.com/bakulskilab).

## Acknowledgments

We thank the participants and staff of the Health and Retirement Study. The Health and Retirement Study is supported by the National Institute on Aging (U01 AG009740).

## Author Contributions

RCH: Data curation, Formal analysis, Visualization, Writing – original draft; HW: Formal analysis, Writing – Review & editing; DJB: Writing – Review & editing; EBW: Conceptualization, Funding acquisition; KMB: Conceptualization, Writing – Review & editing, Funding acquisition

## Conflict of Interest

We have no known conflict of interest to disclose.

## Study Funding

This analysis was supported by the National Institute on Aging (R01 AG067592, R01 AG055406, P30 AG072931)

---

**Supplementary Table 1.**
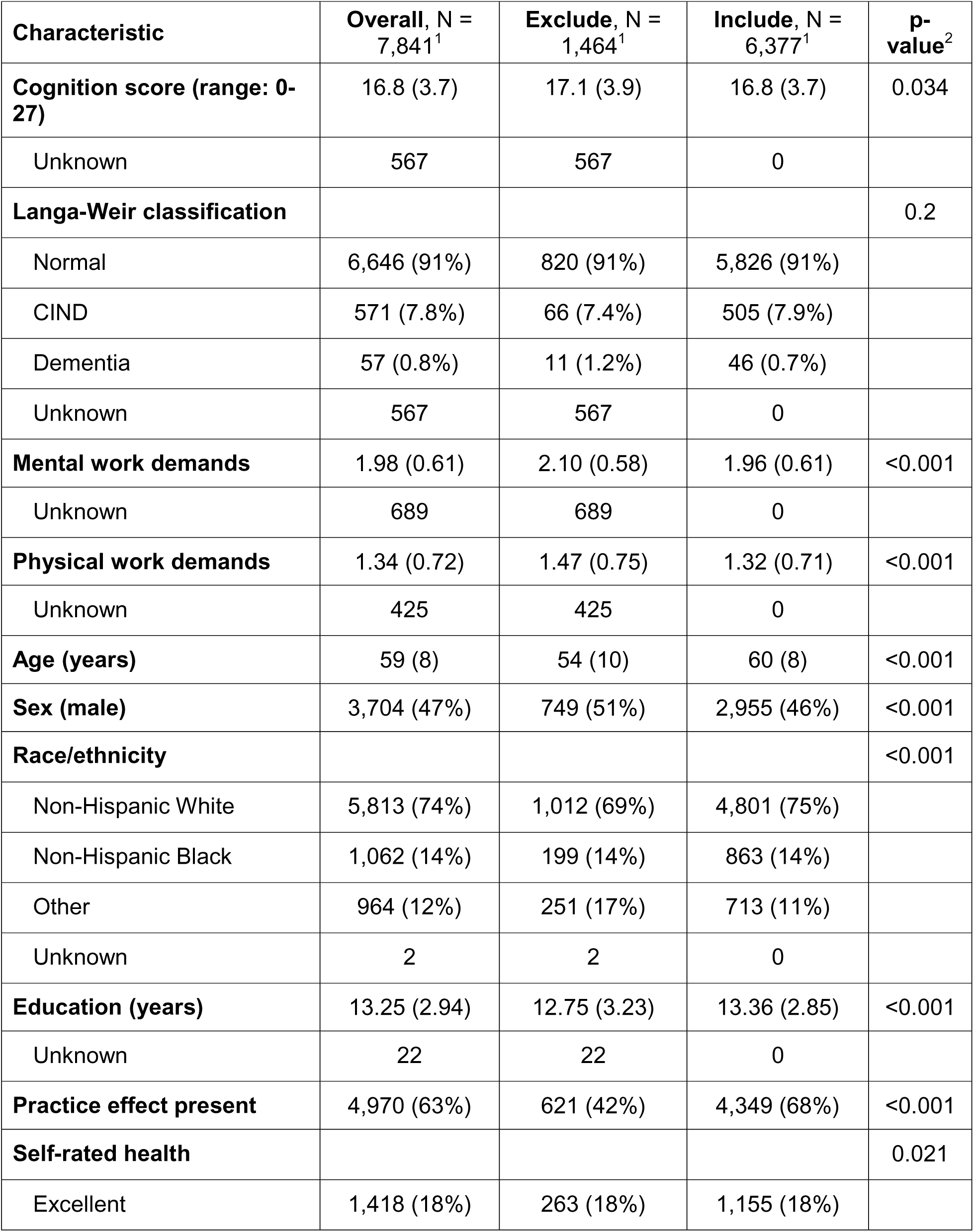
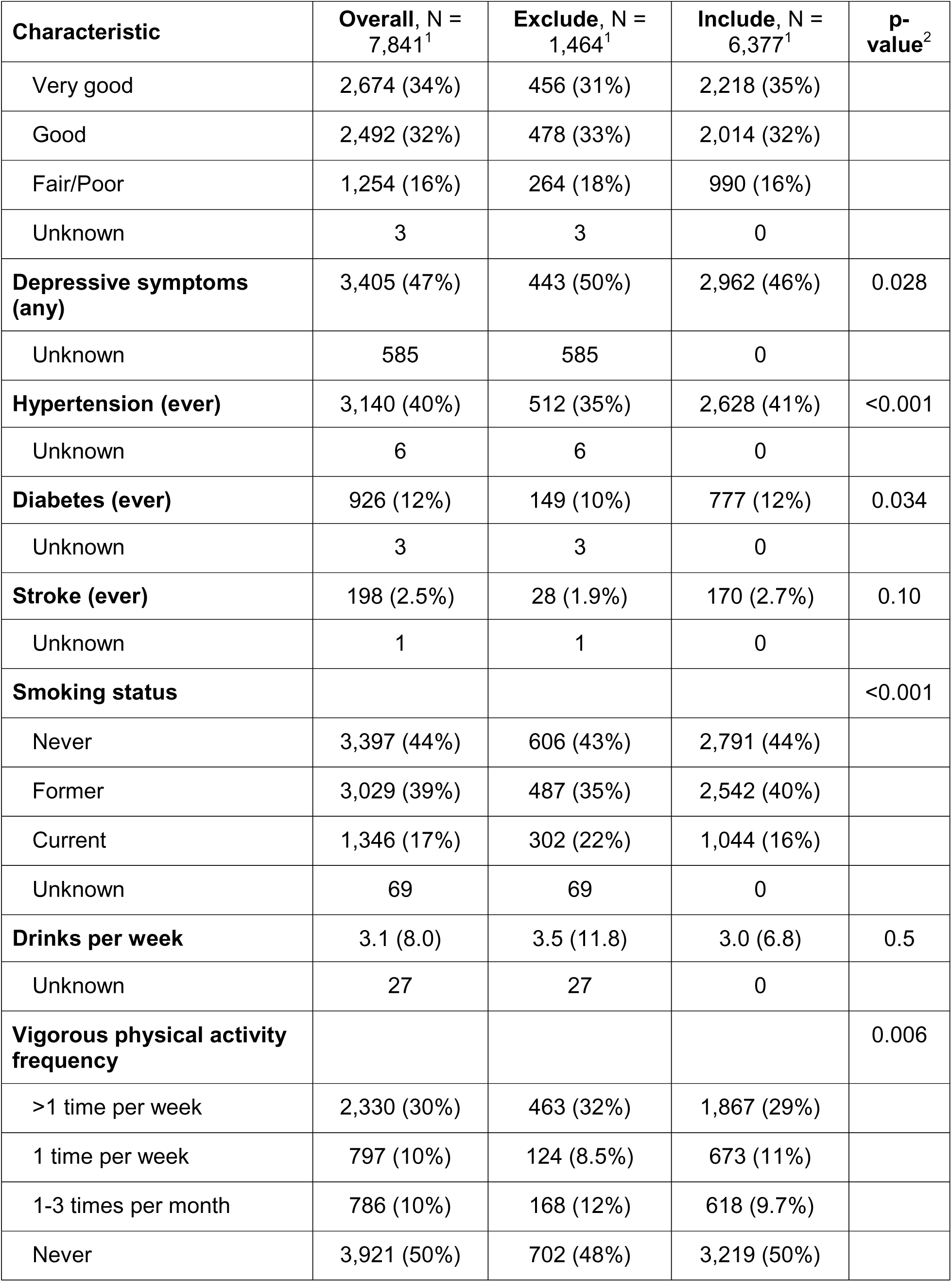
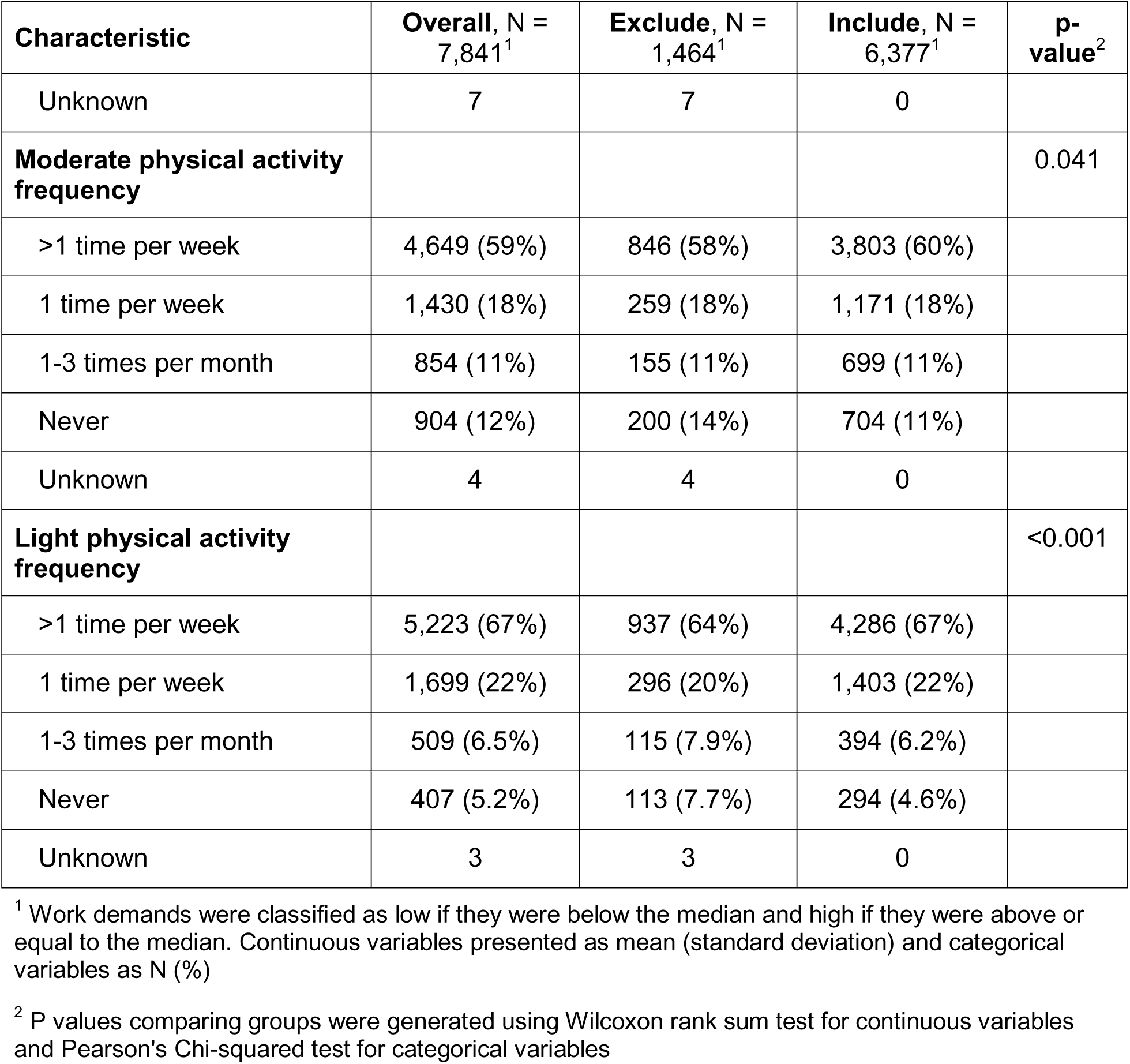
Univariate characteristics of study participants working for pay from the 2004 wave in the Health and Retirement Study, included (n = 6377) vs. excluded (1,464)

**Supplementary Table 2.**
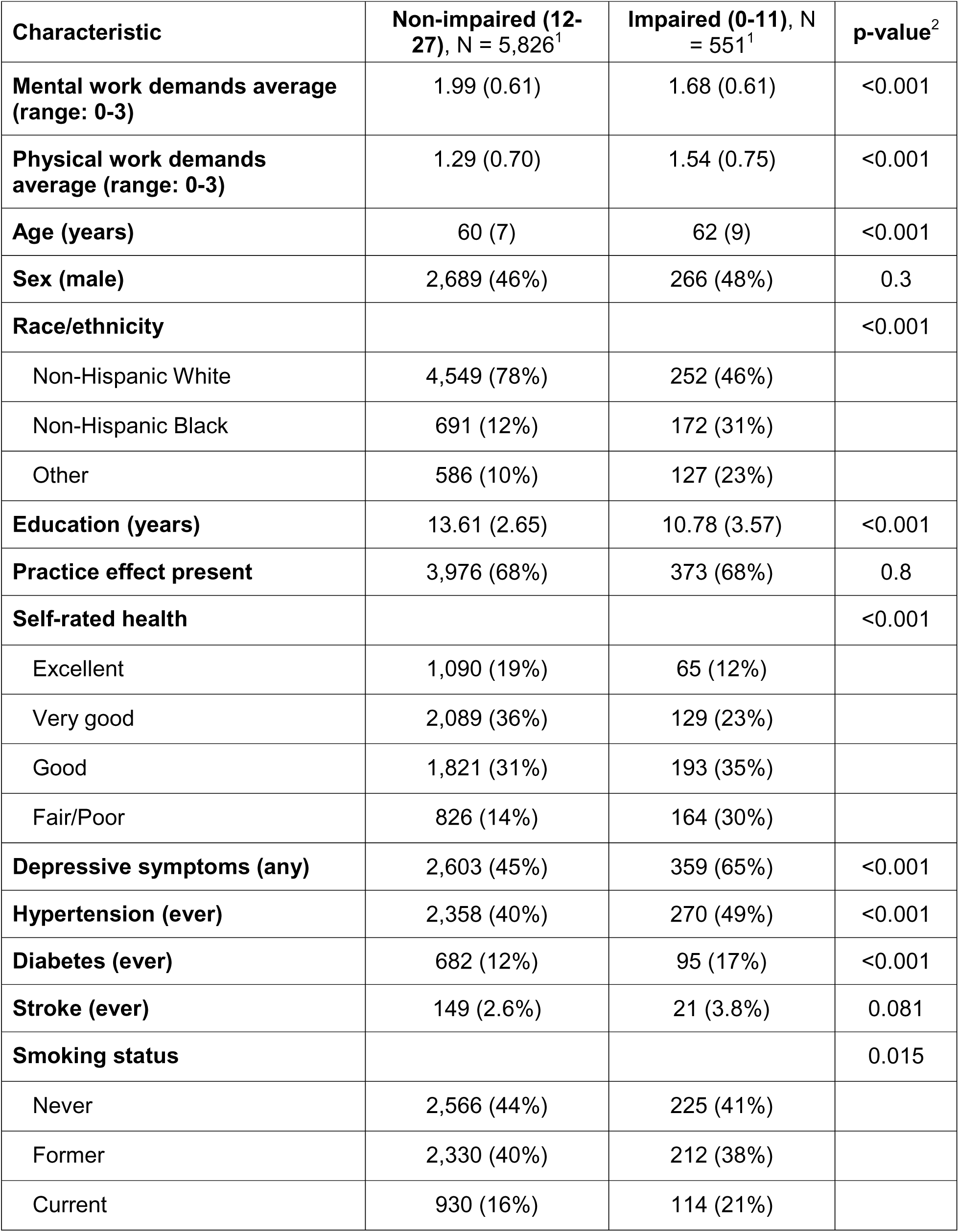
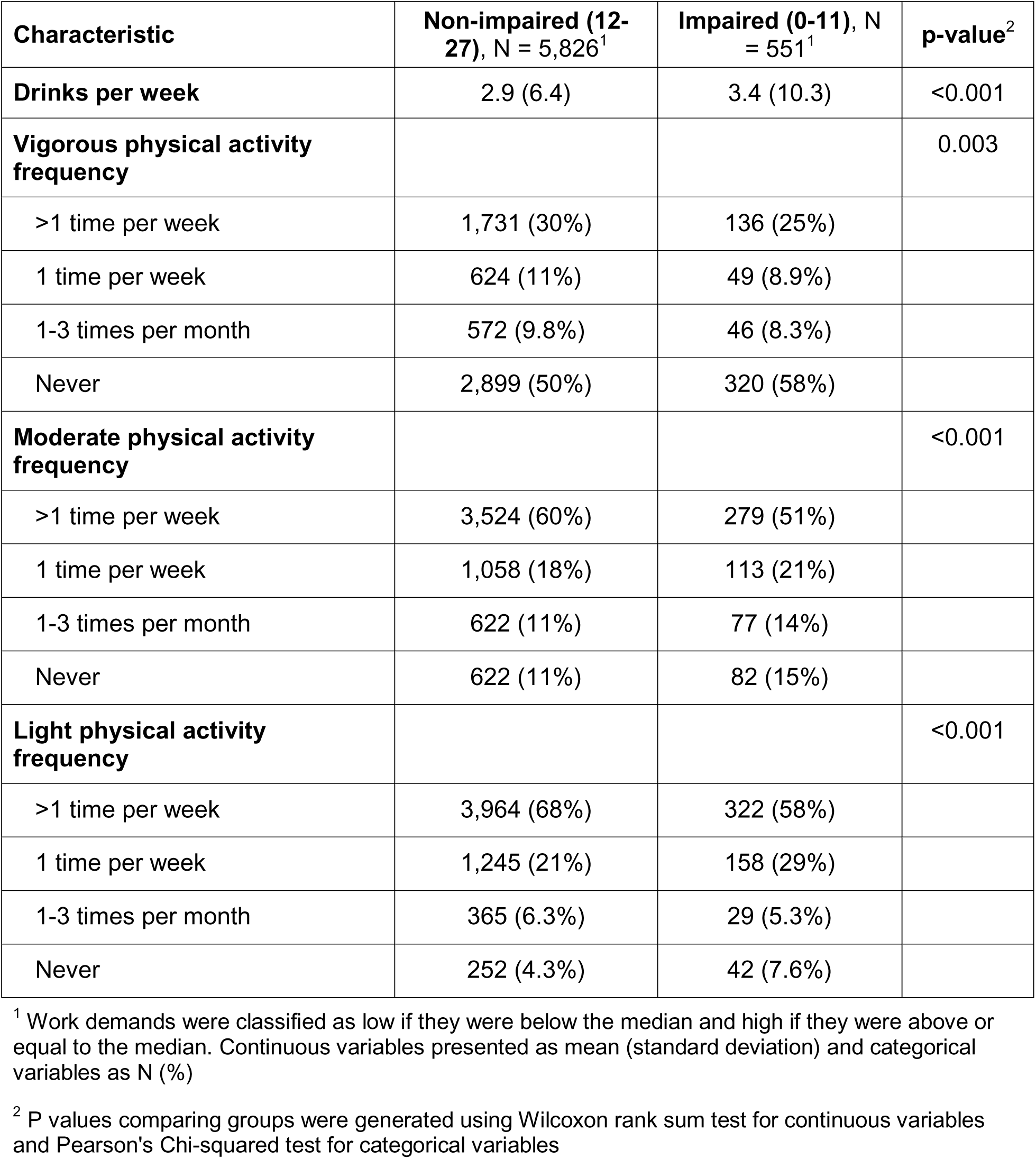
Demographic and occupational characteristics of included study participants by impaired cognition status, Health and Retirement Study, 2004 wave.

| Characteristic | Non-impaired (12-27), N = 5,826<sup>1</sup> | Impaired (0-11), N = 551<sup>1</sup> | p-value<sup>2</sup> |
| --- | --- | --- | --- |
| **Mental work demands average (range: 0-3)** | 1.99 (0.61) | 1.68 (0.61) | <0.001 |
| **Physical work demands average (range: 0-3)** | 1.29 (0.70) | 1.54 (0.75) | <0.001 |
| **Age (years)** | 60 (7) | 62 (9) | <0.001 |
| **Sex (male)** | 2,689 (46%) | 266 (48%) | 0.3 |
| **Race/ethnicity** |  |  | <0.001 |
| Non-Hispanic White | 4,549 (78%) | 252 (46%) |  |
| Non-Hispanic Black | 691 (12%) | 172 (31%) |  |
| Other | 586 (10%) | 127 (23%) |  |
| **Education (years)** | 13.61 (2.65) | 10.78 (3.57) | <0.001 |
| **Practice effect present** | 3,976 (68%) | 373 (68%) | 0.8 |
| **Self-rated health** |  |  | <0.001 |
| Excellent | 1,090 (19%) | 65 (12%) |  |
| Very good | 2,089 (36%) | 129 (23%) |  |
| Good | 1,821 (31%) | 193 (35%) |  |
| Fair/Poor | 826 (14%) | 164 (30%) |  |
| **Depressive symptoms (any)** | 2,603 (45%) | 359 (65%) | <0.001 |
| **Hypertension (ever)** | 2,358 (40%) | 270 (49%) | <0.001 |
| **Diabetes (ever)** | 682 (12%) | 95 (17%) | <0.001 |
| **Stroke (ever)** | 149 (2.6%) | 21 (3.8%) | 0.081 |
| **Smoking status** |  |  | 0.015 |
| Never | 2,566 (44%) | 225 (41%) |  |
| Former | 2,330 (40%) | 212 (38%) |  |
| Current | 930 (16%) | 114 (21%) |  |

| **Characteristic** | **Non-impaired (12-27), N = 5,826<sup>1</sup>** | **Impaired (0-11), N = 551<sup>1</sup>** | **p-value<sup>2</sup>** |
| --- | --- | --- | --- |
| **Drinks per week** | 2.9 (6.4) | 3.4 (10.3) | <0.001 |
| **Vigorous physical activity frequency** |  |  | 0.003 |
| >1 time per week | 1,731 (30%) | 136 (25%) |  |
| 1 time per week | 624 (11%) | 49 (8.9%) |  |
| 1-3 times per month | 572 (9.8%) | 46 (8.3%) |  |
| Never | 2,899 (50%) | 320 (58%) |  |
| **Moderate physical activity frequency** |  |  | <0.001 |
| >1 time per week | 3,524 (60%) | 279 (51%) |  |
| 1 time per week | 1,058 (18%) | 113 (21%) |  |
| 1-3 times per month | 622 (11%) | 77 (14%) |  |
| Never | 622 (11%) | 82 (15%) |  |
| **Light physical activity frequency** |  |  | <0.001 |
| >1 time per week | 3,964 (68%) | 322 (58%) |  |
| 1 time per week | 1,245 (21%) | 158 (29%) |  |
| 1-3 times per month | 365 (6.3%) | 29 (5.3%) |  |
| Never | 252 (4.3%) | 42 (7.6%) |  |

<sup>1</sup> Work demands were classified as low if they were below the median and high if they were above or equal to the median. Continuous variables presented as mean (standard deviation) and categorical variables as N (%)
<sup>2</sup> P values comparing groups were generated using Wilcoxon rank sum test for continuous variables and Pearson's Chi-squared test for categorical variables

**Supplemental Figure 1.**
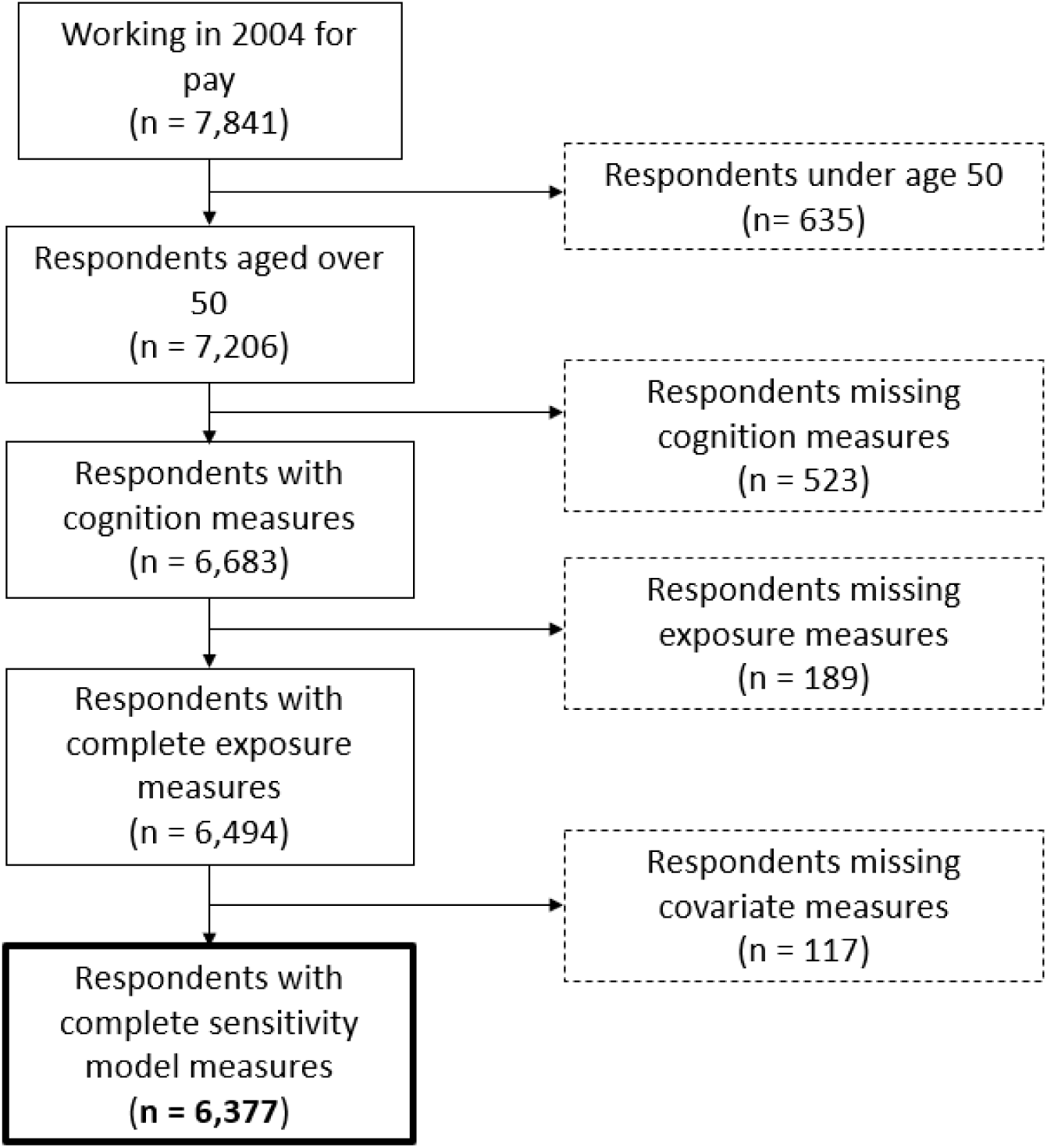
Sample selection steps, Health and Retirement Study, 2004 wave.

## Notes

### Competing Interest Statement

The authors have declared no competing interest.

### Funding Statement

This analysis was funded by the National Institute on Aging (R01 AG067592, R01 AG055406, P30 AG072931)

### Author Declarations

The study used ONLY openly available human data that were originally located at: https://hrsdata.isr.umich.edu/data-products.

